# Race is not Associated with Medical Emergency Team Evaluation Prior to In-Hospital Cardiac Arrest

**DOI:** 10.1101/2024.04.23.24306256

**Authors:** Cody-Aaron L. Gathers, Ryan W. Morgan, Jessica S. Alvey, Ron Reeder, Zachary D. Goldberger, Jessica Fowler, Maryam Y. Naim, Amanda O’Halloran, Raina M. Merchant, Martha Kienzle, Vinay Nadkarni, Robert A. Berg, Robert M. Sutton, the American Heart Association’s Get With The Guidelines®-Resuscitation Investigators

## Abstract

**Background:** Black patients have worse outcomes after in-hospital cardiac arrest (IHCA). Whether these racial disparities are associated with medical emergency team (MET) evaluation prior to IHCA remains unknown.

**Methods:** A retrospective cohort study of adults age ≥ 18 years from the American Heart Association Get With The Guidelines® Resuscitation registry who had an IHCA between 2000 and 2021 with acute physiologic decline (modified early warning score [MEWS] ≥ 3) during the 24 hours prior to IHCA. A propensity-weighted cohort was constructed to balance confounders between Black and White patients. The association between race and MET evaluation was quantified with weighted multivariable logistic regression.

**Results:** Among 354,480 patients, 88,507 met the initial inclusion criteria, of which 29,714 patients (median age 69 [IQR 58-79] years, 42.5% female, and 26.9% Black) had acute physiologic decline during the 24 hours prior to IHCA. Among patients with acute physiologic decline, 4102 (13.8%) patients had a preceding MET evaluation before IHCA. Rates of MET evaluation prior to cardiac arrest did not differ significantly between Black and White patients with acute physiologic decline (aOR 1.02, 95% CI 0.94-1.11, p = 0.62).

**Conclusions:** Though racial disparities in IHCA outcomes exist, this study did not detect a difference in rates of MET evaluation prior to IHCA among patients with acute physiologic decline as a potential mechanism for these disparities.

**Clinical Perspective What Is New?:**

- With the goal to identify potential mechanisms leading to racial disparities in outcomes from adult in-hospital cardiac arrest (IHCA), this multicenter prospective observational study is the first study to analyze the association between race and medical emergency team (MET) evaluation among patients with evidence of clinical deterioration. IHCA
- We found that rates of MET evaluation among patients with clinical deterioration (i.e., vital sign abnormalities) prior to IHCA did not differ between Black and White patients.

**What Are the Clinical Implications?:**

- These data demonstrate that race is not associated with MET evaluation prior to adult in-hospital cardiac arrest.
- Future studies should evaluate other links in the American Heart Association’s in-hospital chain of survival (e.g., resuscitation quality, post-arrest care) as potential targets to mitigate racial disparities in IHCA survival.

## Introduction

Among adults who sustain an in-hospital cardiac arrest (IHCA), Black patients have lower survival rates compared to White patients.^1–3^ Delayed defibrillation, higher severity of chronic medical conditions, and variations in hospital quality have been postulated as potential mechanisms for the racial disparity in IHCA survival.^2,4–6^ However, the reason for this disparity largely remains elusive.

A medical emergency team (MET), also known as a rapid response team, serves to respond to patients at risk of acute decompensation to prevent IHCA and has been integrated as a key link in the American Heart Association (AHA) IHCA Chain of Survival.^7,8^ However, the association between METs and IHCA incidence and in-hospital mortality is inconsistent.^9–11^. A previous study demonstrated that only 17% of patients had a MET evaluation prior to IHCA and most patients without a MET evaluation prior to IHCA had at least one severe vital sign abnormality within 4 hours prior to IHCA.^12^ To our knowledge, prior studies have not evaluated whether racial differences in MET evaluation prior to IHCA exist. Given the well-documented racial disparity in IHCA outcomes and the potential role of a MET in preventing IHCA, an analysis of the role of race as a contributor to MET evaluation prior to IHCA is warranted.

The primary aim of this study was to evaluate the association of race with MET evaluation for patients at risk of acute decompensation prior to IHCA. Acknowledging the documented racial disparities in hospital care, we hypothesized that Black patients would be less likely to receive a MET evaluation prior to IHCA as compared to White patients.^13^

## Methods

### Study population

A retrospective cohort study utilizing the American Heart Association (AHA) Get With The Guidelines®-Resuscitation (GWTG-R) registry was conducted. The GWTG-R program is provided by the AHA. This national registry is a large, voluntary, multi-center, prospective database of IHCA which has been previously described in detail.^14^ Hospitals participating in the registry submit clinical information regarding the medical history, hospital care, and outcomes of consecutive patients hospitalized for cardiac arrest using an online, interactive case report form and Patient Management Tool—PMT™ (IQVIA, Parsippany, New Jersey). All participating institutions were required to comply with local regulatory and privacy guidelines and, if required, to secure institutional review board approval. Because data were used primarily at the local site for quality improvement, sites were granted a waiver of informed consent under the common rule. IQVIA serves as the data collection (through their PMT) and coordination center for the American Heart Association/American Stroke Association GWTG-R programs.

Within the registry, cardiac arrest is defined as either pulselessness or a pulse with inadequate perfusion requiring chest compressions and/or defibrillation of ventricular fibrillation or pulseless ventricular tachycardia. Quality improvement hospital personnel enroll patients with a cardiac arrest without do-not-resuscitate orders who undergo cardiopulmonary resuscitation (CPR). The registry utilizes standardized Utstein-style definitions to report patient variables and outcomes across sites.^15,16^ Data accuracy is ensured by thorough training of research personnel and standardized software. This research did not meet the definition of human subjects research by Children’s Hospital of Philadelphia and was deemed exempt from human subjects research (IRB 21-019305).

### Cohort Design

The study cohort was obtained from hospitals that submitted data on adults 18 years of age or older to the GWTG-R registry between January 1, 2000 and May 10, 2021. The GWTG-R registry cardiopulmonary arrest (CPA) and MET databases were linked and the cohort was derived from hospitals with 10 or more IHCAs that reported both CPA and MET data. Given the primary exposure of Black or White race, this study excluded patients of other races or patients for whom race information was incomplete or missing. The cohort was further confined to only include index IHCAs that occurred after the date of the site’s first MET evaluation as a surrogate of when the MET was instituted at a given hospital. We further excluded patients with a cardiac arrest not on a general inpatient unit (defined as general inpatient area, rehab, skilled nursing, or mental health unit) or a telemetry unit (defined as adult coronary unit, telemetry unit, or step-down unit) as MET evaluations do not typically occur in these locations. Similarly to a previous study analyzing missed opportunities for MET evaluations, we further delineated the cohort to include patients who had a MET evaluation on a general inpatient unit or telemetry unit, transferred to the ICU after MET evaluation, and afterward had an IHCA within 24 hours of transfer to assign credit for the MET evaluation.^12^ The primary cohort was additionally restricted to exclude patients with missing vital signs or a modified early warning score (MEWS) < 3.

### Study variables and outcomes

The primary exposure variable was race (Black vs. White) among patients with acute physiologic decline (MEWS ≥ 3) within 24 hours of cardiac arrest. Race was classified as identified in the patient’s medical record. The primary outcome was MET evaluation (Yes vs. No). Patient data included demographic characteristics (age, sex), illness category (medical cardiac, medical non-cardiac, surgical-cardiac, surgical non-cardiac), cardiac arrest location (general inpatient unit, telemetry unit, or ICU), pre-existing conditions (acute central nervous system [CNS] non-stroke event; acute stroke; baseline CNS depression; congestive heart failure [current or prior]; diabetes mellitus; renal, respiratory, or hepatic insufficiency; hypotension or hypoperfusion; major trauma; metastatic or hematologic malignancy; metabolic or electrolyte abnormality; myocardial infarction [current or prior]; sepsis or pneumonia); initial rhythm of cardiac arrest (ventricular fibrillation, pulseless ventricular tachycardia, asystole, pulseless electrical activity, bradycardia, or other/unknown), interventions in place before the cardiac arrest (antiarrhythmic infusions, arterial catheter, dialysis, electrocardiogram monitor, supplemental oxygen, vascular access, vasoactive agent), whether the event was witnessed, timing of the cardiac arrest (weekday [7:00 AM-10:59 PM], weeknight [11:00 PM-6:59 AM], weekend, or holiday), event duration, medications administered during arrest (epinephrine, vasopressors other than epinephrine bolus, sodium bicarbonate, calcium chloride or gluconate, atropine, lidocaine, amiodarone), and MEWS.

Given that vital sign abnormalities may signify that a patient warranted a MET evaluation, we included patients with a MEWS ≥ 3 within 24 hours of the IHCA. The MEWS is a bedside evaluation tool based on five physiologic parameters: systolic blood pressure, heart rate, respiratory rate, temperature, and level of consciousness for a maximum score of 14.^17^ Specific values for assignment of the MEWS are outlined in Supplemental Table 1. A MEWS > 4 signifies a high risk for death.^18^ For this analysis, MEWS was calculated as a composite of blood pressure, heart rate, respiratory rate, and temperature for a maximum MEWS of 11 (level of consciousness is not available in the database). As METs serve to intervene on patients before cardiac arrest (i.e., before they are at high risk of death), we incorporated a more sensitive cut off for MEWS (≥ 3), which may signify acute physiologic decline and consideration for MET evaluation.

Hospital characteristics included hospital teaching status, hospital bed number, geographic region, and hospital racial composition. Hospital teaching status was defined as major teaching (fellowship program), minor teaching (only a residency program), or nonteaching (no residency or fellowship program). Hospital bed number was categorized as less than 200 beds, 200-499 beds, or 500 or more beds. Geographic region was characterized as North Mid Atlantic, South Atlantic and Puerto Rico, North Central, South Central, or Mountain and Pacific. Hospital racial composition was defined as a hospital with a patient population that is less than 10% Black, 10%-30% Black, or more than 30% Black adopted from a previous study analyzing hospital racial composition and cardiac arrest outcomes.^6^ Hospital racial composition has been shown to influence cardiac arrest outcomes with hospitals with a higher proportion of Black patients having worse outcomes after cardiac arrest.^6^

### Statistical analysis

To assess the association of race with MET evaluation, propensity weighted regression was utilized to reduce potential confounding due to a priori selected variables including age, sex, illness category, pre-existing conditions, cardiac arrest location, and hospital characteristics.^19^ While propensity weighted regression is traditionally used to balance confounders between treatment groups, it is equally well-suited to balance confounders between Black and White patients. Three steps were utilized to construct a cohort of Black and White patients that are balanced with respect to potentially confounding factors.

First, a multivariable logistic regression model of race was created using the variables described above as predictors. For patients with any missing pre-existing condition, “missing” was coded as a separate category to indicate the patient was missing this information and to equally balance missing data between the propensity-matched cohorts. Variables included in the propensity model included age, sex, illness category, pre-existing conditions, cardiac arrest location, and hospital characteristics. A butterfly plot of propensities was constructed for Black and White patients with acute physiologic decline within 24 hours of cardiac arrest (Supplemental Figure 1). Patients with propensity scores outside of the region of overlap between the histograms were excluded from the final multivariable model assessing the association between race and MET evaluation.

Second, patients were weighted utilizing stabilized inverse probability of treatment weights. This weighting increases the balance of a priori characteristics between Black and White patients, creating a weighted cohort in which patients differ with respect to race but are balanced with respect to potentially confounding variables. This is the mechanism by which confounding from characteristics is reduced. Differences in means and proportions were compared between Black and White patients. The cohorts were balanced when the absolute standardized difference for each variable was <0.10 (Supplemental Table 2).

Third, a logistic regression model was constructed using the weighted cohort to assess the association of race and MET evaluation prior to IHCA for all patients with acute physiologic decline. The model utilized a robust variance estimator to account for the correlation due to clustering of patients within hospitals and further adjusted for hospital teaching status to account for variability of MET implementation across hospitals.^12^

All analyses were performed using SAS 9.4 (SAS Institute; Cary, NC) and the AHA Precision Medicine Platform (https://precision.heart.org/) with reported p-values based on a two-sided alternative and considered significant if less than 0.05.

## Results

Among 354,480 patients at eligible hospitals, 29,714 who met inclusion criteria had acute physiologic decline within 24 hours prior to IHCA (Figure 1). Baseline characteristics of the balanced cohorts are included in Table 1. The median age was 69 (IQR 58-79) years, 12,643 (42.5%) were female, and 7993 (26.9%) were Black. The median MEWS was 4 (IQR 3-5), and vital signs were measured at a median time of 114 minutes (IQR 58-202) prior to IHCA.

**Figure 1:**
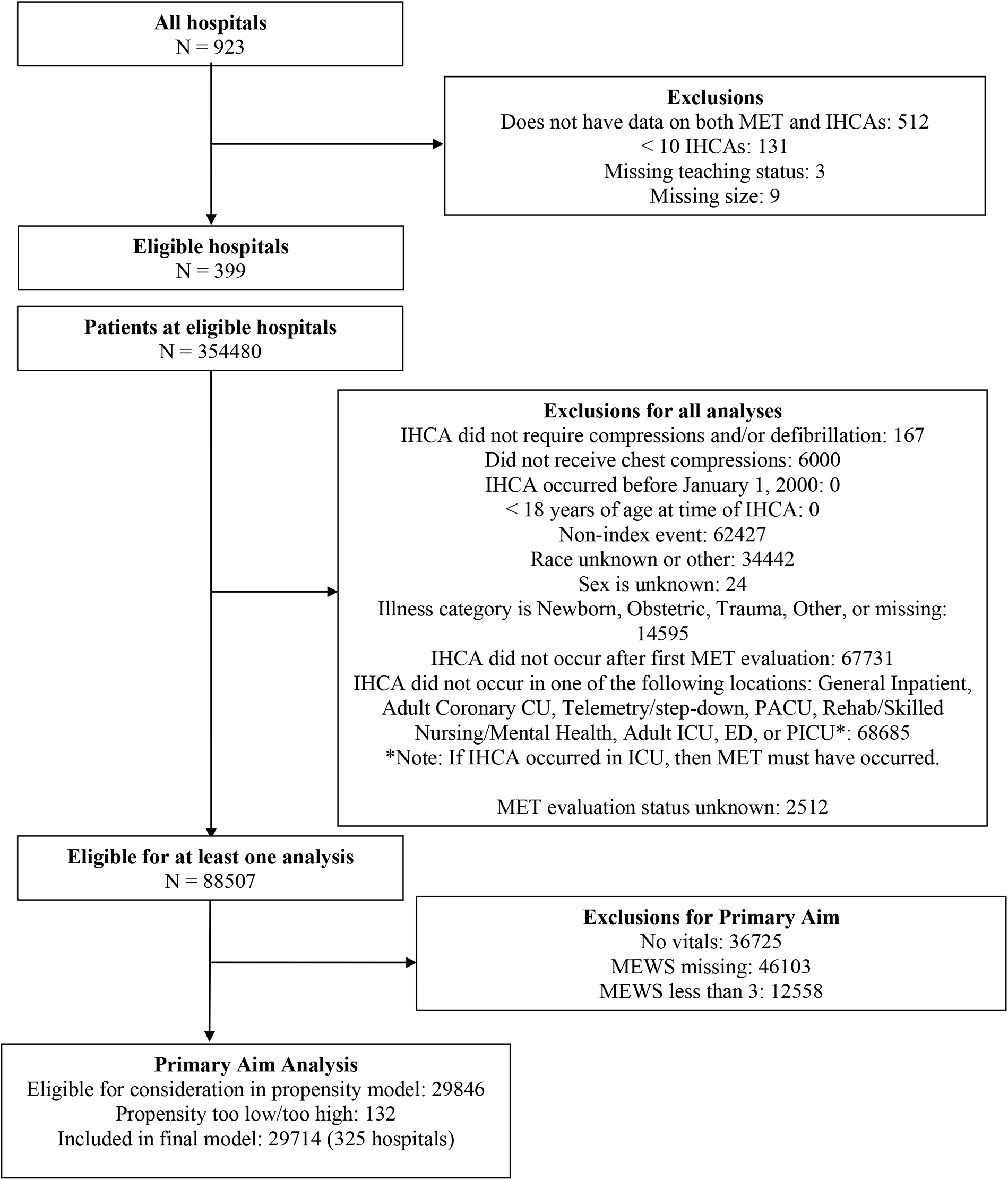
CONSORT Diagram

**Table 1:** Primary Cohort Characteristics (Acute Physiologic Decline)

|  | **White**<br>(N = 21721) | **Black**<br>(N = 7993) | **Overall**<br>(N = 29714) |
| --- | --- | --- | --- |
| **Patient and Event Characteristics** |  |  |  |
| Age (years) | 71.0 [60.0, 80.0] | 64.0 [54.0, 74.0] | 69.0 [58.0, 79.0] |
| Sex |  |  |  |
| Male | 12928 (59.5%) | 4143 (51.8%) | 17071 (57.5%) |
| Female | 8793 (40.5%) | 3850 (48.2%) | 12643 (42.5%) |
| Illness category |  |  |  |
| Medical-cardiac | 8026 (37.0%) | 2450 (30.7%) | 10476 (35.3%) |
| Medical-noncardiac | 10127 (46.6%) | 4579 (57.3%) | 14706 (49.5%) |
| Surgical-cardiac | 1100 (5.1%) | 215 (2.7%) | 1315 (4.4%) |
| Surgical-noncardiac | 2468 (11.4%) | 749 (9.4%) | 3217 (10.8%) |
| Location of cardiac arrest |  |  |  |
| General Inpatient | 8448 (38.9%) | 3269 (40.9%) | 11717 (39.4%) |
| Telemetry Unit | 12089 (55.7%) | 4214 (52.7%) | 16303 (54.9%) |
| Intensive Care Unit | 1184 (5.5%) | 510 (6.4%) | 1694 (5.7%) |
| Event witnessed | 15979 (73.6%) | 5765 (72.1%) | 21744 (73.2%) |
| Night, weekend, or holiday event | 11712 (53.9%) | 4254 (53.2%) | 15966 (53.7%) |
| MEWS before cardiac arrest | 4.0 [3.0, 5.0] | 4.0 [3.0, 5.0] | 4.0 [3.0, 5.0] |
| MEWS time relative to cardiac arrest (minutes) | 113.0<br>[57.0, 201.0] | 115.0<br>[60.0, 202.0] | 114.0<br>[58.0, 202.0] |
| **Pre-Existing Conditions** |  |  |  |
| Missing pre-existing condition | 2282 (10.5%) | 768 (9.6%) | 3050 (10.3%) |
| Acute CNS non-stroke event | 1219 (5.6%) | 583 (7.3%) | 1802 (6.1%) |
| Acute stroke | 635 (2.9%) | 322 (4.0%) | 957 (3.2%) |
| Baseline depression in CNS function | 1705 (7.8%) | 774 (9.7%) | 2479 (8.3%) |
| Congestive heart failure (this admission) | 3887 (17.9%) | 1340 (16.8%) | 5227 (17.6%) |
| Congestive heart failure (prior to this admission) | 5188 (23.9%) | 2019 (25.3%) | 7207 (24.3%) |
| Diabetes mellitus | 6860 (31.6%) | 2934 (36.7%) | 9794 (33.0%) |
| Hepatic insufficiency | 1484 (6.8%) | 628 (7.9%) | 2112 (7.1%) |
| Hypotension/Hypoperfusion | 3833 (17.6%) | 1439 (18.0%) | 5272 (17.7%) |
| Major trauma | 215 (1.0%) | 60 (0.8%) | 275 (0.9%) |
| Metastatic or hematologic malignancy | 2808 (12.9%) | 1150 (14.4%) | 3958 (13.3%) |
| Metabolic/electrolyte abnormality | 3512 (16.2%) | 1461 (18.3%) | 4973 (16.7%) |
| Myocardial ischemia/infarction (this admission) | 2716 (12.5%) | 633 (7.9%) | 3349 (11.3%) |

**Table 1: Primary Cohort Characteristics (Acute Physiologic Decline)**
|  |  |  |  |
| --- | --- | --- | --- |
| Myocardial ischemia/infarction (prior to this admission) | 3500 (16.1%) | 729 (9.1%) | 4229 (14.2%) |
| Renal insufficiency | 6785 (31.2%) | 3386 (42.4%) | 10171 (34.2%) |
| Respiratory insufficiency | 7573 (34.9%) | 2686 (33.6%) | 10259 (34.5%) |
| Sepsis/pneumonia (including septicemia) | 5338 (24.6%) | 2057 (25.7%) | 7395 (24.9%) |
| **Interventions** |  |  |  |
| Antiarrhythmic infusions | 403 (1.9%) | 75 (0.9%) | 478 (1.6%) |
| Arterial catheter | 1100 (5.1%) | 440 (5.5%) | 1540 (5.2%) |
| Dialysis | 422 (1.9%) | 272 (3.4%) | 694 (2.3%) |
| ECG monitor | 15964 (73.6%) | 5661 (70.9%) | 21625 (72.9%) |
| Supplemental oxygen | 9587 (44.1%) | 3290 (41.2%) | 12877 (43.3%) |
| Vascular access | 20101 (92.7%) | 7316 (91.7%) | 27417 (92.4%) |
| Vasoactive agent | 2973 (13.7%) | 1089 (13.6%) | 4062 (13.7%) |
| **Arrest Characteristics** |  |  |  |
| First documented rhythm |  |  |  |
| Asystole or PEA | 15903 (73.2%) | 6260 (78.3%) | 22163 (74.6%) |
| Ventricular fibrillation or pulseless ventricular tachycardia | 2929 (13.5%) | 719 (9.0%) | 3648 (12.3%) |
| Bradycardia | 952 (4.4%) | 404 (5.1%) | 1356 (4.6%) |
| Other or unknown | 1937 (8.9%) | 610 (7.6%) | 2547 (8.6%) |
| CPR duration (minutes)1 | 16.0 [8.0, 27.0] | 16.0 [8.0, 27.0] | 16.0 [8.0, 27.0] |
| Epinephrine bolus | 19086 (87.9%) | 7331 (91.7%) | 26417 (88.9%) |
| Total number of epinephrine doses | 3.0 [2.0, 5.0] | 3.0 [2.0, 5.0] | 3.0 [2.0, 5.0] |
| Time to first epinephrine (minutes)2 | 2.0 [1.0, 4.0] | 2.0 [1.0, 4.0] | 2.0 [1.0, 4.0] |
| Epinephrine dosing interval (minutes) | 6.0 [4.2, 9.0] | 5.8 [4.0, 8.5] | 6.0 [4.2, 8.8] |
| Vasopressor(s) other than epinephrine bolus | 6121 (28.2%) | 2386 (29.9%) | 8507 (28.6%) |
| Sodium bicarbonate | 11617 (53.5%) | 4838 (60.5%) | 16455 (55.4%) |
| Calcium chloride or calcium gluconate | 6261 (28.8%) | 2845 (35.6%) | 9106 (30.6%) |
| Atropine | 8929 (41.1%) | 3344 (41.8%) | 12273 (41.3%) |
| Lidocaine | 930 (4.3%) | 299 (3.7%) | 1229 (4.1%) |
| Amiodarone | 3889 (17.9%) | 1263 (15.8%) | 5152 (17.3%) |
| **Hospital characteristics** |  |  |  |
| Teaching hospital |  |  |  |
| None | 2819 (13.0%) | 697 (8.7%) | 3516 (11.8%) |
| Minor | 5992 (27.6%) | 4120 (51.5%) | 10112 (34.0%) |
| Major | 12910 (59.4%) | 3176 (39.7%) | 16086 (54.1%) |

**Table 1: Primary Cohort Characteristics (Acute Physiologic Decline)**
|  |  |  |  |
| --- | --- | --- | --- |
| Hospital size |  |  |  |
| Less than 200 beds | 2290 (10.5%) | 575 (7.2%) | 2865 (9.6%) |
| 200 - 499 beds | 10644 (49.0%) | 2907 (36.4%) | 13551 (45.6%) |
| 500 or more beds | 8787 (40.5%) | 4511 (56.4%) | 13298 (44.8%) |
| Site region |  |  |  |
| North Mid Atlantic | 2381 (11.0%) | 742 (9.3%) | 3123 (10.5%) |
| South Atlantic and Puerto Rico | 6127 (28.2%) | 2908 (36.4%) | 9035 (30.4%) |
| North Central | 5440 (25.0%) | 1420 (17.8%) | 6860 (23.1%) |
| South Central | 5416 (24.9%) | 2738 (34.3%) | 8154 (27.4%) |
| Mountain and Pacific | 2357 (10.9%) | 185 (2.3%) | 2542 (8.6%) |
| Site Racial Composition |  |  |  |
| Less than 10% Black | 7840 (36.1%) | 401 (5.0%) | 8241 (27.7%) |
| 10-30% Black | 8083 (37.2%) | 2191 (27.4%) | 10274 (34.6%) |
| More than 30% Black | 5798 (26.7%) | 5401 (67.6%) | 11199 (37.7%) |

<sup>1</sup>Truncated after 120 minutes
<sup>2</sup>Truncated after 30 minutes
CPA = cardiopulmonary arrest, MEWS = modified early warning score, CNS = central nervous system, ECG = electrocardiogram, PEA = pulseless electrical activity, CPR = cardiopulmonary resuscitation

Among patients with acute physiologic decline, 4102 (13.8%) had a documented MET evaluation prior to IHCA. After adjusting for potential confounders, there was no significant difference in MET evaluation between Black vs. White patients with acute physiologic decline within 24 hours prior to IHCA (aOR 1.02, 95% CI 0.94-1.11, p = 0.62, Table 2).

**Table 2:** Primary Cohort Association of Race with MET Evaluation (acute physiologic decline)

|  | MET Evaluation |  |
| --- | --- | --- |
|  | Odds ratio<br>(95% CI) | P-value |
| **Black** |  | 0.617 |
| White | Reference |  |
| Black | 1.02 (0.94, 1.11) |  |
| **Teaching hospital** |  | 0.795 |
| Major | 0.94 (0.69, 1.27) |  |
| Minor | Reference |  |
| None | 1.04 (0.70, 1.55) |  |

Output shows estimates based on patient race. Model adjusts for hospital teaching status. The model also uses a robust variance estimator to account for within-hospital correlation, and weights patients according to their propensity.

## Discussion

In this large, multi-center, propensity weighted cohort study of MET evaluation prior to IHCA, we failed to detect an association between race and MET evaluation prior to IHCA. This is notable as racial disparities in IHCA outcomes do not appear to be attributed to differences in rates of MET evaluations between Black and White patients.

Previous studies have shown racial differences in cardiac arrest outcomes.^1–3^ While some studies suggest that delayed defibrillation, severity of chronic illnesses, and variation in hospital quality may contribute to the racial disparities in IHCA outcomes, no study has convincingly identified a mechanism for these disparities.^2,4,5^ A previous study demonstrated that only 1 out of 7 patients has a MET evaluation prior to IHCA, and many patients have severely abnormal vital signs prior to IHCA.^12^ Given the low rate of MET evaluation prior to IHCA and the role or MET evaluation in potentially mitigating IHCA, our study builds upon previous observations by evaluating the association of race and MET evaluation prior to IHCA to better elucidate a potential mechanism for racial disparities in IHCA outcomes.

The lack of an association between race and MET evaluation prior to IHCA can potentially be explained by patient acuity prior to cardiac arrest—a patient who is acutely decompensating should warrant a MET evaluation and ICU transfer regardless of their race. While this study did not identify a statistically significant relationship between race and MET evaluation prior to IHCA, it is important to continue to recognize the role of implicit bias in clinical decision making, particularly in high acuity environments where implicit bias may be magnified.^20–22^ Additionally, there may be competing needs (notifying other medical teams or goals of care discussions with families) prior to cardiac arrest, and a MET evaluation might not be prioritized in that moment.^12^ Moreover, the composition and implementation of METs is subject to significant variability between hospitals.^12,23^ This variability further contributes to a lack of standardization for MET intervention and may further explain the lack of an association between race and MET evaluation prior to IHCA. Our study further adjusted for hospital teaching status to potentially account for this variability and found no statistically significant difference in the odds of MET evaluation between Black and White patients.

Given that we did not detect an association between race and MET evaluations prior to cardiac arrest, the results of this study suggest that further investigation is required to further clarify the mechanisms for disparate IHCA outcomes between Black and White patients. Early recognition and prevention of cardiac arrest is one link in the AHA IHCA Chain of Survival.^8^ Further research should investigate and further clarify the contribution of high-quality CPR, defibrillation, post-cardiac arrest care, and recovery to improve these core domains of the AHA Chain of Survival in hospital resuscitation systems. Given this study and the known differences in hospital resuscitation systems, particular consideration on racial disparities in high-quality CPR and post-cardiac arrest care can be prioritized.^24,25^

This study must be interpreted in the context of its limitations. The data available in GWTG-R do not account for unmeasured confounders that may contribute to MET evaluation. Notable potential confounders that were not collected that are associated with adverse outcomes include socioeconomic status, the primary language of the patient, the presence of a family member at the bedside, hospital staffing, hospital acuity at the time of the IHCA, and experience of the primary team.^12,26–30^ Additionally, there is significant variability in the composition and implementation of METs between hospitals that cannot be completely accounted for by hospital teaching status.^12^ Moreover, the hospitals that participate in GWTG-R may have more robust MET evaluation systems in place than other hospitals, potentially limiting generalizability. We are unable to determine if there was a delay in timing of MET evaluation in relation to the first detection of acute physiologic decline, an important factor known to affect patient outcomes after MET evaluation. Lastly, there is a potential for exposure misclassification as race is collected from the medical record.

## Conclusion

In this propensity weighted cohort study, no association between race and MET evaluation prior to IHCA was detected as a potential mechanism for racial disparities in IHCA outcomes. Further investigation is required to examine mechanisms behind racial disparities in IHCA outcomes.

## Data Availability

In accordance with the journal's data sharing policy, all data supporting the findings of this study are available within the article and its supplementary materials.

## Acknowledgements

We would like to acknowledge the GTWG-R Adult Research Task Force: Anne Grossestreuer PhD; Ari Moskowitz MD MPH; Joseph Ornato MD FACP FACC FACEP; Matthew Churpek MD MPH PhD; Monique Anderson Starks MD MHS; Paul Chan MD MSc; Saket Girotra MD SM; Sarah Perman MD MSCE

## Abbreviations

IHCA: In-Hospital Cardiac Arrest
MET: Medical Emergency Team
AHA: American Heart Association
ICU: Intensive Care Unit
GWTG-R: Get With The Guidelines-Resuscitation
PMT: Patient Management Tool
CPR: Cardiopulmonary Resuscitation
CPA: Cardiopulmonary Arrest
MEWS: Modified Early Warning Score
CNS: Central Nervous System
ROC: Return Of Circulation
ROSC: Return Of Spontaneous Circulation
CPC: Cerebral Performance Category
aOR: Adjusted Odds Ratio

## Authorship contribution statement

Cody-Aaron Gathers: conceptualization, data interpretation, drafting of the original manuscript, critical revision of the manuscript

Ryan Morgan: conceptualization, data interpretation, critical revision of the manuscript, supervision

Jessica Alvey: conceptualization, statistical analysis, data interpretation, critical revision of the manuscript

Ron Reeder: conceptualization, statistical analysis, data interpretation, critical revision of the manuscript

Zachary Goldberger: critical revision of the manuscript

Jessica Fowler: conceptualization, critical revision of the manuscript

Maryam Naim: conceptualization, critical revision of the manuscript

Amanda O’Halloran: conceptualization, critical revision of the manuscript

Raina Merchant: conceptualization, critical revision of the manuscript

Martha Kienzle: conceptualization, critical revision of the manuscript

Vinay Nadkarni: conceptualization, critical revision of the manuscript

Robert Berg: conceptualization, critical revision of the manuscript

Bobby Sutton: conceptualization, data interpretation, critical revision of the manuscript, supervision

